# Virtual care use prior to emergency department admissions during a stable COVID-19 period in Ontario, Canada

**DOI:** 10.1101/2022.10.20.22281298

**Authors:** V. Stamenova, C. Chu, E. Borgundvaag, C. Fleury, J. Brual, O. Bhattacharyya, M. Tadrous

**Affiliations:** Women’s College Hospital; ICES; University of Toronto Leslie Dan Faculty of Pharmacy

## Abstract

**Background:** The increased use of telemedicine during the pandemic has led to concerns about potential increased emergency department (ED) admissions and outpatient service use prior to such admissions. We examined the frequency of telemedicine use prior to ED admissions and characterized the patients with prior telemedicine use and the physicians who provided these outpatient visits.

**Methods:** We conducted a retrospective, population-based, cross-sectional analysis using linked health administrative data in Ontario, Canada to identify patients who had an ED admission between July 1 and September 30, 2021 and patients with an ED admissions during the same period in 2019. We grouped patients based on their use of outpatient services in the 7 days prior to admission and reported their sociodemographic characteristics and healthcare utilization.

**Results:** There were 1,080,334 ED admissions in 2021 vs. 1,113,230 in 2019. In 2021, 74% of these admissions had no prior outpatient visits (virtual or in-person) within 7 days of admission, compared to 75% in 2019. Only 3% of ED admissions had both virtual and in-person visits in the 7 days prior to ED admission. Patients with prior virtual care use were more likely to be hospitalized than those without any outpatient care (13% vs 7.7.%).

**Interpretation:** The net amount of ED admissions and outpatient care prior to admission remained the same over a period of the COVID-19 pandemic when cases were relatively stable. Virtual care seems to be able to appropriately triage patients to the ED and may even prove beneficial for diverting patients away from the ED when an ED admission is not appropriate.

The COVID-19 pandemic has led to the emergence of standard use of telemedicine in health care across the globe(1,2). In Ontario, Canada the proportion of ambulatory visits completed virtually has been maintained at slightly above 50% from 2020 to 2021 (3). Despite its widespread adoption, it is still unclear when virtual visits are clinically appropriate and how such wide use of telemedicine impacts patient outcomes and healthcare utilization metrics.

Before the pandemic, there had been concerns that telemedicine may lead to an increased use of outpatient services with patients having both a virtual and an in-person visit for the same clinical issue(4,5). For example, pre-pandemic data (2007-2016) from Manitoba showed that telemedicine users had on average 1.3 times more ambulatory visits than non-users.(6) In addition, studies have produced mixed evidence with regard to the effect of telemedicine on urgent services such as emergency department (ED) admissions and hospitalizations (7). Many of the studies reported in the literature are based on data from site-specific programs and therefore have limited generalizability. Finally, policymakers and some physicians have become concerned that the high rates of telemedicine during COVID-19 have led to an increase in emergency department admissions because of poor access to in-person outpatient care (8). This concern is exacerbated when one considers rural and lower socioeconomic status patients who already had poor access to care before the pandemic(9). Combined with reports of lower uptake of telemedicine among these patients(10,11), it is not clear how the transition of care from in-person to virtual impacts ED use.

The high adoption of telemedicine during the pandemic, in the context of a publicly funded healthcare system allowing us access to most visits across the entire population, offers a unique opportunity to examine the frequency of telemedicine use prior to ED admissions. Therefore, the goal of this study was to characterize the frequency and modality (in-person vs virtual) of outpatient care prior to ED admissions. We examined whether there was an overall increase in outpatient visits prior to ED admissions during a period of the pandemic when access to telemedicine was available compared to a seasonality matched period before the pandemic where access to telemedicine was quite limited.

We also aimed to characterize the patients who had a telemedicine visit prior to an ED admission vs. those who had an in-person visit and the physicians who saw patients with virtual only visits prior to their ED admission compared to those who saw patients virtually or in-person prior to their ED admission.

## Methods

### Study Design and Population

We conducted a retrospective, population-based, cross-sectional analysis using linked health administrative data in Ontario, Canada to identify all patients who had an ED admission between July 1 and September 30, 2021 and those with ED admissions between July 1 and September 30, 2019. The 2021 summer window represented a relatively stable period of COVID-19, in between the major waves of COVID-19 infection and the 2019 period served as a control period during which access to telemedicine was relatively limited. We excluded patients who had invalid identification numbers, were non-Ontario residents, had ED admissions that were not publicly insured, and those who had another ED admission within 7 days prior to July 1 of the year of interest (full exclusion list is provided in the supplement).

### Data sources

Ontario is the province with the largest population in Canada and all permanent residents have public insurance fully covering all necessary physician and hospital services. We used the Ontario Health Insurance Plan (OHIP) for physician claims, the Canadian Institutes of Health Information Discharge Abstract Database (CIHI-DAD) for information about all hospitalizations, the CIHI National Ambulatory Care Reporting System (NACRS) for hospital- and community-based ambulatory care including ED admissions, and the ICES Physician Database (IPDB) for data on physician specialty. Databases were linked using unique identifiers and analyzed at ICES (formerly the Institute for Clinical Evaluative Sciences). Virtual visits were identified as any OHIP claim with the location recorded as “P” for phone, indicating virtual/telemedicine services. We then described patients based on characteristics, such as age, sex, chronic disease diagnoses, income quintile (based on postal code), urban vs. rural (based on postal code’ s rurality index for Ontario (RIO) where a score below 40 was categorized as urban), Ontario Marginalization Index (ONMARG) containing data on patient economic, ethno-racial, age-based, and social deprivation (details on databases used in the supplement).

### Patient Groups

Patients were categorized into subgroups based on the type of outpatient visit (in-person vs virtual) they had prior to their ED admission (if any). For patients who had virtual visits prior to their ED admission (within 7 days), subgroups of patients were also created based on whether their last virtual outpatient visit was within 24, 48, 72 hours, or 7 days of their ED admission. The following non-mutually exclusive groups of patients were created:

1. all patients with an ED admission during the study window
2. patients with an ED admission who did <u>not</u> have any virtual visits within the 7 days prior (includes patients who had zero visits or only in-person visits prior to ED admission)
3. patients with an ED admission who did not have any outpatient visits (virtual or in-person) within the 7 days prior
4. patients who had at least one virtual visit within 24 hours prior to an ED admission
5. patients who had at least one virtual visit within 48 hours prior to an ED admission
6. patients who had at least one virtual visit within 72 hours prior to an ED admission
7. patients who had at least one virtual visit within 7 days prior to an ED admission
8. patients who had only in-person visits during the 7 days prior to admission.

Patients may have belonged to more than one subgroup, i.e. the 48 hours virtual care group includes patients in the 24 hours group. All analyses were conducted using SAS version 9.4.

### Patient and physician characteristics

To compare the characteristics of patients who had a virtual vs. an in-person visit prior to ED admission, we looked at the most recent visit that occurred within 7 days prior to their ED admission. We identified patient characteristics such as age, sex, region of residence, rurality, neighborhood income, marginalization index, chronic conditions, number of ED admissions, hospitalizations and outpatient visits in the year prior to ED admission, number of outpatient visits in the 7 days prior to ED admission, days between outpatient visit and emergency visit, whether the visit was on the same day as the ED admission, and if the ED admission resulted in hospitalization.

To examine characteristics of the outpatient visits that occurred within 7 days before ED admissions, we classified patients into 4 groups. Three of these groups included patients who had visited the ED in 2021: those with any visit (virtual or in-person), those with a virtual visit, and those with an in-person visit prior to ED admission. The last group consisted of patients who had an in-person visit prior to ED admission in 2019 (we did not include a virtual visit group as virtual visits were uncommon in 2019).

We also looked at the characteristics of physicians who had provided the most recent outpatient visit prior to the patient’ s ED admission in 2021 and categorized them into two groups based on the type of visit: physicians who had seen patients with only a virtual visit in 2021; physicians who had seen patients with either a virtual or in-person visit in 2021. For physician characteristics, we looked at age, sex, region of and years in practice, and patient daily volume.

See Supplement for detailed definitions.

### Ethics

Use of these databases for the purposes of this study was authorized under §45 of Ontario’ s Personal Health Information Protection Act, which does not require review by a research ethics board (REB). All data was de-identified at ICES and individual patient consent was waived.

## Results

Between July 1, 2021 and September 30, 2021, there were 1,080,334 ED admissions in Ontario and 74% of these admissions had no prior outpatient visits (virtual or in-person) within 7 days of admission (Figure 1). Furthermore, 14% of patients admitted to ED had at least one virtual visit within the 7 days prior to ED admission, occurring on average 2.25 (SD=2.31) days before the ED admission. Among those who had a virtual visit within 7 days of ED admission, 34% had a virtual visit on the same day as the ED admission. (Table 1.1) In comparison, between July 1, 2019 and September 30, 2019, there were 1,113,230 ED admissions and 75% of admissions had no prior outpatient visits (virtual or in-person) within 7 days of admission and only 0.7% of admissions had a virtual visit 7 days prior to ED admission. (Table 1.2)

**Table 1.1.** Sociodemographic characteristics of patients with ED admissions in July 1, 2021-September 30, 2021..

|  | All patients | No prior virtual visits | No prior visits (virtual or in-person) | Virtual Visits within 24 hours | Virtual Visits within 48 hours | Virtual Visits within 72 hours | Virtual Visits within 7 days | In-person Visits only within 7 days |
| --- | --- | --- | --- | --- | --- | --- | --- | --- |
|  | N=1,080,334 | N=929,179 | N=802,433 | N=76,027 | N=92,749 | N=106,801 | N=151,155 | N=126,746 |
| **Age** |  |  |  |  |  |  |  |  |
| <18 | 180,889<br>(16.74%) | 162,735<br>(17.51%) | 144,582<br>(18.02%) | 12,073<br>(15.88%) | 13,731 (14.8%) | 14,900<br>(13.95%) | 18,154<br>(12.01%) | 18,153<br>(14.32%) |
| 18-34 | 241,155<br>(22.32%) | 210,946<br>(22.7%) | 187,122<br>(23.32%) | 15,378<br>(20.23%) | 18,799<br>(20.27%) | 21,624<br>(20.25%) | 30,209<br>(19.99%) | 23,824 (18.8%) |
| 35-49 | 185,769<br>(17.2%) | 158,539<br>(17.06%) | 139,117<br>(17.34%) | 13,423<br>(17.66%) | 16,529<br>(17.82%) | 19,085<br>(17.87%) | 27,230<br>(18.01%) | 19,422<br>(15.32%) |
| 50-64 | 204,831<br>(18.96%) | 174,337<br>(18.76%) | 150,059<br>(18.7%) | 14,386<br>(18.92%) | 17,858<br>(19.25%) | 20,891<br>(19.56%) | 30,494<br>(20.17%) | 24,278<br>(19.15%) |
| 65+ | 267,690<br>(24.78%) | 222,622<br>(23.96%) | 181,553<br>(22.63%) | 20,767<br>(27.32%) | 25,832<br>(27.85%) | 30,301<br>(28.37%) | 45,068<br>(29.82%) | 41,069 (32.4%) |
| **Sex** |  |  |  |  |  |  |  |  |
| Female | 563,375<br>(52.15%) | 474,322<br>(51.05%) | 404,643<br>(50.43%) | 44,354<br>(58.34%) | 54,326<br>(58.57%) | 62,631<br>(58.64%) | 89,053<br>(58.92%) | 69,679<br>(54.98%) |
| Male | 516,959<br>(47.85%) | 454,857<br>(48.95%) | 397,790<br>(49.57%) | 31,673<br>(41.66%) | 38,423<br>(41.43%) | 44,170<br>(41.36%) | 62,102<br>(41.08%) | 57,067<br>(45.02%) |
| **Region** |  |  |  |  |  |  |  |  |
| Central | 304,291<br>(28.17%) | 246,835<br>(26.56%) | 207,806<br>(25.9%) | 30,796<br>(40.51%) | 37,046<br>(39.94%) | 42,144<br>(39.46%) | 57,456<br>(38.01%) | 39,029<br>(30.79%) |
| Central East | 275,906<br>(25.54%) | 239,363<br>(25.76%) | 209,307<br>(26.08%) | 17,725<br>(23.31%) | 21,848<br>(23.56%) | 25,292<br>(23.68%) | 36,543<br>(24.18%) | 30,056<br>(23.71%) |
| North | 93,309 (8.64%) | 87,084 (9.37%) | 79,109 (9.86%) | 2,483 (3.27%) | 3,240 (3.49%) | 3,895 (3.65%) | 6,225 (4.12%) | 7,975 (6.29%) |
| Toronto Central | 77,225 (7.15%) | 62,678 (6.75%) | 52,603 (6.56%) | 7,657 (10.07%) | 9,242 (9.96%) | 10,566 (9.89%) | 14,547 (9.62%) | 10,075 (7.95%) |
| West | 329,603<br>(30.51%) | 293,219<br>(31.56%) | 253,608<br>(31.6%) | 17,366<br>(22.84%) | 21,373<br>(23.04%) | 24,904<br>(23.32%) | 36,384<br>(24.07%) | 39,611<br>(31.25%) |
| **Residence** |  |  |  |  |  |  |  |  |
| Rural | 120,461<br>(11.15%) | 112,309<br>(12.09%) | 103,599<br>(12.91%) | 3,298 (4.34%) | 4,252 (4.58%) | 5,123 (4.8%) | 8,152 (5.39%) | 8,710 (6.87%) |
| Urban | 939,612<br>(86.97%) | 797,861<br>(85.87%) | 681,515<br>(84.93%) | 72,175<br>(94.93%) | 87,787<br>(94.65%) | 100,849<br>(94.43%) | 141,751<br>(93.78%) | 116,346<br>(91.79%) |
| **Missing** | 20,261 (1.88%) | 19,009 (2.05%) | 17,319 (2.16%) | 554 (0.73%) | 710 (0.77%) | 829 (0.78%) | 1,252 (0.83%) | 1,690 (1.33%) |
| **Neighbourhood income quintile** |  |  |  |  |  |  |  |  |
| **1 (lowest)** | 245,567 (22.73%) | 213,754 (23%) | 184,360 (22.98%) | 15,226 (20.03%) | 18,711 (20.17%) | 21,834 (20.44%) | 31,813 (21.05%) | 29,394 (23.19%) |
| **2** | 219,408 (20.31%) | 188,441 (20.28%) | 162,383 (20.24%) | 15,281 (20.1%) | 18,752 (20.22%) | 21,648 (20.27%) | 30,967 (20.49%) | 26,058 (20.56%) |
| **3** | 214,567 (19.86%) | 184,130 (19.82%) | 158,999 (19.81%) | 15,576 (20.49%) | 19,011 (20.5%) | 21,804 (20.42%) | 30,437 (20.14%) | 25,131 (19.83%) |
| **4** | 207,048 (19.17%) | 177,256 (19.08%) | 153,432 (19.12%) | 15,222 (20.02%) | 18,538 (19.99%) | 21,258 (19.9%) | 29,792 (19.71%) | 23,824 (18.8%) |
| **5 (highest)** | 189,830 (17.57%) | 162,155 (17.45%) | 140,370 (17.49%) | 14,471 (19.03%) | 17,428 (18.79%) | 19,906 (18.64%) | 27,675 (18.31%) | 21,785 (17.19%) |
| **Missing** | 3,914 (0.36%) | 3,443 (0.37%) | 2,889 (0.36%) | 251 (0.33%) | 309 (0.33%) | 351 (0.33%) | 471 (0.31%) | 554 (0.44%) |
| **Marginalization** |  |  |  |  |  |  |  |  |
| **Dependency** | 2.95 ± 1.48 | 2.98 ± 1.48 | 2.99 ± 1.48 | 2.69 ± 1.47 | 2.71 ± 1.47 | 2.73 ± 1.47 | 2.77 ± 1.48 | 2.91 ± 1.49 |
| **Material deprivation** | 3.04 ± 1.43 | 3.06 ± 1.43 | 3.06 ± 1.43 | 2.90 ± 1.43 | 2.91 ± 1.43 | 2.92 ± 1.43 | 2.94 ± 1.44 | 3.04 ± 1.44 |
| **Residential instability** | 3.13 ± 1.43 | 3.13 ± 1.42 | 3.13 ± 1.42 | 3.03 ± 1.51 | 3.04 ± 1.50 | 3.05 ± 1.50 | 3.09 ± 1.49 | 3.16 ± 1.46 |
| **Ethnic concentration** | 3.05 ± 1.47 | 2.99 ± 1.46 | 2.94 ± 1.46 | 3.55 ± 1.38 | 3.53 ± 1.39 | 3.51 ± 1.39 | 3.45 ± 1.41 | 3.26 ± 1.44 |
| **Asthma** | 203,600 (18.85%) | 171,869 (18.5%) | 147,570 (18.39%) | 15,224 (20.02%) | 18,768 (20.24%) | 21,810 (20.42%) | 31,731 (20.99%) | 24,299 (19.17%) |
| **CHF** | 37,196 (3.44%) | 30,305 (3.26%) | 23,964 (2.99%) | 3,039 (4%) | 3,804 (4.1%) | 4,521 (4.23%) | 6,891 (4.56%) | 6,341 (5%) |
| **COPD** | 44,642 (4.13%) | 36,727 (3.95%) | 30,029 (3.74%) | 3,384 (4.45%) | 4,276 (4.61%) | 5,123 (4.8%) | 7,915 (5.24%) | 6,698 (5.28%) |
| **Dementia** | 22,035 (2.04%) | 18,700 (2.01%) | 14,551 (1.81%) | 1,631 (2.15%) | 1,997 (2.15%) | 2,316 (2.17%) | 3,335 (2.21%) | 4,149 (3.27%) |
| **HIV** | 2,403 (0.22%) | 1,996 (0.21%) | 1,649 (0.21%) | 159 (0.21%) | 205 (0.22%) | 259 (0.24%) | 407 (0.27%) | 347 (0.27%) |
| **Hypertension** | 299,654 (27.74%) | 247,745 (26.66%) | 203,248 (25.33%) | 23,581 (31.02%) | 29,595 (31.91%) | 34,765 (32.55%) | 51,909 (34.34%) | 44,497 (35.11%) |
| **Crohn's** | 10,369 (0.96%) | 8,344 (0.9%) | 6,987 (0.87%) | 895 (1.18%) | 1,139 (1.23%) | 1,330 (1.25%) | 2,025 (1.34%) | 1,357 (1.07%) |
| **Diabetes** | 162,984 (15.09%) | 132,973 (14.31%) | 108,000 (13.46%) | 13,518 (17.78%) | 16,958 (18.28%) | 19,976 (18.7%) | 30,011 (19.85%) | 24,973 (19.7%) |
| **Arthritis** | 15,908 (1.47%) | 12,762 (1.37%) | 10,233 (1.28%) | 1,373 (1.81%) | 1,726 (1.86%) | 2,046 (1.92%) | 3,146 (2.08%) | 2,529 (2%) |
| **ED visits in past 365d** | 2.46 ± 3.62 | 2.43 ± 3.59 | 2.38 ± 3.42 | 2.40 ± 3.14 | 2.48 ± 3.44 | 2.53 ± 3.48 | 2.65 ± 3.78 | 2.69 ± 4.42 |
| **Hospitalizations in past 365d** | 1.48 ± 1.15 | 1.47 ± 1.14 | 1.45 ± 1.12 | 1.47 ± 1.06 | 1.49 ± 1.07 | 1.50 ± 1.09 | 1.55 ± 1.18 | 1.52 ± 1.19 |
| **Physician visits in past 365d** | 12.56 ± 18.24 | 11.38 ± 16.91 | 10.30 ± 15.24 | 17.11 ± 22.34 | 17.68 ± 22.70 | 18.09 ± 22.89 | 19.04 ± 23.26 | 17.54 ± 23.41 |
| **Number of outpatient visits in past 7d** | 1.32 ± 0.65 | 1.17 ± 0.46 | . ± . | 1.52 ± 0.82 | 1.51 ± 0.81 | 1.50 ± 0.80 | 1.45 ± 0.76 | 1.17 ± 0.46 |
| **Number of days between virtual/in-person and ED visits** | 2.25 ± 2.31 | . ± . | . ± . | 0.32 ± 0.47 | 0.63 ± 0.77 | 0.94 ± 1.08 | 2.25 ± 2.31 | 2.55 ± 2.47 |
| **Virtual/in-person visit same day as ED visit, N (%)** | 51,448 (4.76%) | 0 (0.00%) | 0 (0.00%) | 51,448 (67.67%) | 51,448 (55.47%) | 51,448 (48.17%) | 51,448 (34.04%) | 42,845 (33.8%) |
| **ED visit resulted in hospitalization, N (%)** | 98,735 (9.14%) | 79,860 (8.59%) | 61,462 (7.66%) | 8,874 (11.67%) | 11,025 (11.89%) | 12,897 (12.08%) | 18,875 (12.49%) | 18,398 (14.52%) |

**Table 1.2.** Sociodemographic characteristics of patients with ED admissions in July 1, 2019-September 30, 2019.

|  | All patients | No prior virtual visits | No prior visits (virtual or in-person) | Virtual Visits within 24 hours | Virtual Visits within 48 hours | Virtual Visits within 72 hours | Virtual Visits within 7 days | In-person Visits only within 7 days |
| --- | --- | --- | --- | --- | --- | --- | --- | --- |
|  | N=1,113,230 | N=1,105,823 | N=833,255 | N=3,313 | N=4,196 | N=4,974 | N=7,407 | N=272,568 |
| **Age, N (%)** |  |  |  |  |  |  |  |  |
| <18 | 195,715 (17.58%) | 195,142 (17.65%) | 155,250 (18.63%) | 339 (10.23%) | 421 (10.03%) | 469 (9.43%) | 573 (7.74%) | 39,892 (14.64%) |
| 18-34 | 248,772 (22.35%) | 246,597 (22.3%) | 194,330 (23.32%) | 927 (27.98%) | 1,195 (28.48%) | 1,435 (28.85%) | 2,175 (29.36%) | 52,267 (19.18%) |
| 35-49 | 189,026 (16.98%) | 187,167 (16.93%) | 143,137 (17.18%) | 708 (21.37%) | 948 (22.59%) | 1,164 (23.4%) | 1,859 (25.1%) | 44,030 (16.15%) |
| 50-64 | 213,850 (19.21%) | 212,375 (19.21%) | 158,197 (18.99%) | 663 (20.01%) | 824 (19.64%) | 980 (19.7%) | 1,475 (19.91%) | 54,178 (19.88%) |
| 65+ | 265,867 (23.88%) | 264,542 (23.92%) | 182,341 (21.88%) | 676 (20.4%) | 808 (19.26%) | 926 (18.62%) | 1,325 (17.89%) | 82,201 (30.16%) |
| **Sex, N (%)** |  |  |  |  |  |  |  |  |
| Female | 582,163 (52.29%) | 578,305 (52.3%) | 424,729 (50.97%) | 1,740 (52.52%) | 2,194 (52.29%) | 2,593 (52.13%) | 3,858 (52.09%) | 153,576 (56.34%) |
| Male | 531,067 (47.71%) | 527,518 (47.7%) | 408,526 (49.03%) | 1,573 (47.48%) | 2,002 (47.71%) | 2,381 (47.87%) | 3,549 (47.91%) | 118,992 (43.66%) |
| Region, N (%) |  |  |  |  |  |  |  |  |
| Central | 314,480 (28.25%) | 313,156 (28.32%) | 216,889 (26.03%) | 593 (17.9%) | 753 (17.95%) | 877 (17.63%) | 1,324 (17.87%) | 96,267 (35.32%) |
| Central East | 277,966 (24.97%) | 275,813 (24.94%) | 211,043 (25.33%) | 1,023 (30.88%) | 1,273 (30.34%) | 1,506 (30.28%) | 2,153 (29.07%) | 64,770 (23.76%) |
| North | 103,609 (9.31%) | 102,386 (9.26%) | 88,305 (10.6%) | 439 (13.25%) | 590 (14.06%) | 724 (14.56%) | 1,223 (16.51%) | 14,081 (5.17%) |
| Toronto Central | 80,876 (7.26%) | 80,567 (7.29%) | 55,592 (6.67%) | 146 (4.41%) | 182 (4.34%) | 216 (4.34%) | 309 (4.17%) | 24,975 (9.16%) |
| West | 336,299 (30.21%) | 333,901 (30.19%) | 261,426 (31.37%) | 1,112 (33.56%) | 1,398 (33.32%) | 1,651 (33.19%) | 2,398 (32.37%) | 72,475 (26.59%) |
| Residence, N (%) |  |  |  |  |  |  |  |  |
| Rural | 130,792 (11.75%) | 129,847 (11.74%) | 113,488 (13.62%) | 387 (11.68%) | 494 (11.77%) | 598 (12.02%) | 945 (12.76%) | 16,359 (6%) |
| Urban | 960,154 (86.25%) | 953,944 (86.27%) | 700,633 (84.08%) | 2,834 (85.54%) | 3,582 (85.37%) | 4,227 (84.98%) | 6,210 (83.84%) | 253,311 (92.93%) |
| Missing | 22,284 (2%) | 22,032 (1.99%) | 19,134 (2.3%) | 92 (2.78%) | 120 (2.86%) | 149 (3%) | 252 (3.4%) | 2,898 (1.06%) |
| Neighbourhood income quintile, N (%) |  |  |  |  |  |  |  |  |
| 1 (lowest) | 260,878 (23.43%) | 258,723 (23.4%) | 196,750 (23.61%) | 890 (26.86%) | 1,147 (27.34%) | 1,376 (27.66%) | 2,155 (29.09%) | 61,973 (22.74%) |
| 2 | 229,374 (20.6%) | 227,808 (20.6%) | 171,634 (20.6%) | 691 (20.86%) | 879 (20.95%) | 1,039 (20.89%) | 1,566 (21.14%) | 56,174 (20.61%) |
| 3 | 222,006 (19.94%) | 220,722 (19.96%) | 165,857 (19.9%) | 591 (17.84%) | 752 (17.92%) | 898 (18.05%) | 1,284 (17.33%) | 54,865 (20.13%) |
| 4 | 206,948 (18.59%) | 205,631 (18.6%) | 154,243 (18.51%) | 614 (18.53%) | 761 (18.14%) | 889 (17.87%) | 1,317 (17.78%) | 51,388 (18.85%) |
| 5 (highest) | 190,182 (17.08%) | 189,128 (17.1%) | 141,931 (17.03%) | 508 (15.33%) | 638 (15.2%) | 750 (15.08%) | 1,054 (14.23%) | 47,197 (17.32%) |
| Missing | 3,842 (0.35%) | 3,811 (0.34%) | 2,840 (0.34%) | 19 (0.57%) | 19 (0.45%) | 22 (0.44%) | 31 (0.42%) | 971 (0.36%) |
| Marginalization |  |  |  |  |  |  |  |  |
| Dependency | 2.97 ± 1.48 | 2.97 ± 1.48 | 3.02 ± 1.48 | 3.23 ± 1.44 | 3.23 ± 1.44 | 3.24 ± 1.44 | 3.26 ± 1.44 | 2.83 ± 1.48 |
| Material deprivation | 3.09 ± 1.43 | 3.09 ± 1.43 | 3.11 ± 1.43 | 3.26 ± 1.42 | 3.29 ± 1.42 | 3.30 ± 1.41 | 3.35 ± 1.40 | 3.02 ± 1.44 |
| Residential instability | 3.14 ± 1.43 | 3.14 ± 1.43 | 3.14 ± 1.41 | 3.31 ± 1.36 | 3.33 ± 1.36 | 3.33 ± 1.35 | 3.37 ± 1.35 | 3.13 ± 1.48 |
| **Ethnic concentration** | 3.04 ± 1.47 | 3.05 ± 1.47 | 2.93 ± 1.47 | 2.64 ± 1.35 | 2.64 ± 1.36 | 2.64 ± 1.35 | 2.62 ± 1.35 | 3.40 ± 1.43 |
| **Asthma** | 222,572 (19.99%) | 220,756 (19.96%) | 162,633 (19.52%) | 745 (22.49%) | 985 (23.47%) | 1,182 (23.76%) | 1,816 (24.52%) | 58,123 (21.32%) |
| **CHF** | 51,895 (4.66%) | 51,631 (4.67%) | 34,088 (4.09%) | 119 (3.59%) | 144 (3.43%) | 167 (3.36%) | 264 (3.56%) | 17,543 (6.44%) |
| **COPD** | 55,734 (5.01%) | 55,247 (5%) | 37,937 (4.55%) | 182 (5.49%) | 246 (5.86%) | 292 (5.87%) | 487 (6.57%) | 17,310 (6.35%) |
| **Dementia** | 34,090 (3.06%) | 33,978 (3.07%) | 23,174 (2.78%) | 58 (1.75%) | 72 (1.72%) | 83 (1.67%) | 112 (1.51%) | 10,804 (3.96%) |
| **HIV** | 2,474 (0.22%) | 2,439 (0.22%) | 1,647 (0.2%) | 17 (0.51%) | 21 (0.5%) | 24 (0.48%) | 35 (0.47%) | 792 (0.29%) |
| **Hypertension** | 327,408 (29.41%) | 325,435 (29.43%) | 224,901 (26.99%) | 927 (27.98%) | 1,144 (27.26%) | 1,341 (26.96%) | 1,973 (26.64%) | 100,534 (36.88%) |
| **Crohn's** | 11,413 (1.03%) | 11,327 (1.02%) | 7,766 (0.93%) | 33 (1%) | 51 (1.22%) | 58 (1.17%) | 86 (1.16%) | 3,561 (1.31%) |
| **Diabetes** | 169,310 (15.21%) | 168,211 (15.21%) | 114,459 (13.74%) | 485 (14.64%) | 596 (14.2%) | 714 (14.35%) | 1,099 (14.84%) | 53,752 (19.72%) |
| **Arthritis** | 17,520 (1.57%) | 17,382 (1.57%) | 11,507 (1.38%) | 59 (1.78%) | 70 (1.67%) | 89 (1.79%) | 138 (1.86%) | 5,875 (2.16%) |
| **ED visits in past 365d** | 2.53 ± 3.57 | 2.52 ± 3.56 | 2.47 ± 3.46 | 2.99 ± 3.94 | 3.11 ± 4.21 | 3.20 ± 4.36 | 3.37 ± 4.44 | 2.68 ± 3.85 |
| **Hospitalizations in past 365d** | 1.50 ± 1.14 | 1.50 ± 1.14 | 1.48 ± 1.11 | 1.55 ± 1.29 | 1.57 ± 1.30 | 1.60 ± 1.27 | 1.63 ± 1.28 | 1.53 ± 1.18 |
| **Physician visits in past 365d** | 11.68 ± 16.84 | 11.62 ± 16.75 | 9.96 ± 14.48 | 16.46 ± 21.73 | 17.66 ± 23.11 | 18.48 ± 23.21 | 20.70 ± 24.44 | 16.33 ± 21.25 |
| **Number of outpatient visits in past 7d** | 1.25 ± 0.56 | 1.25 ± 0.56 | . ± . | 1.35 ± 0.70 | 1.35 ± 0.70 | 1.35 ± 0.70 | 1.34 ± 0.68 | 1.25 ± 0.56 |
| **Number of days between virtual/in-person and ED visits** | 2.49 ± 2.34 | . ± . | . ± . | 0.33 ± 0.47 | 0.68 ± 0.80 | 1.05 ± 1.12 | 2.49 ± 2.34 | 2.80 ± 2.46 |
| **Virtual/in-person visit same day as ED visit, N (%)** | 2,211 (0.2%) | 0 (0.00%) | 0 (0.00%) | 2,211 (66.74%) | 2,211 (52.69%) | 2,211 (44.45%) | 2,211 (29.85%) | 76,054 (27.9%) |
| **ED visit resulted in hospitalization, N (%)** | 94,415 (8.48%) | 93,747 (8.48%) | 59,781 (7.17%) | 345 (10.41%) | 411 (9.8%) | 466 (9.37%) | 668 (9.02%) | 33,966 (12.46%) |

**Table 2:** Top 5 Reasons for ED admission, N (%), July 1, 2021-September 30, 2021

|  | All patients | No prior virtual visits | No prior visits (virtual or in-person) | Virtual Visits within 24 hours | Virtual Visits within 48 hours | Virtual Visits within 72 hours | Virtual Visits within 7 days | In-person Visits only within 7 days |
| --- | --- | --- | --- | --- | --- | --- | --- | --- |
|  | N=1,080,334 | N=929,179 | N=802,433 | N=76,027 | N=92,749 | N=106,801 | N=151,155 | N=126,746 |
| **Chest pain** | 38,183<br>(14.1%) | 31,859<br>(13.8%) | 27,735<br>(13.6%) | 3,321<br>(15.7%) | 3,964<br>(15.6%) | 4,497<br>(15.5%) | 6,324<br>(15.8%) | 4,124<br>(15.1%) |
| **Abdominal pain** | 29,859<br>(11.0%) | 24,688<br>(10.7%) | 21,029<br>(10.3%) | 2,480<br>(11.8%) | 3,066<br>(12.1%) | 3,536<br>(12.2%) | 5,171<br>(12.9%) | 3,659<br>(13.4%) |
| **Urinary tract infection** | 25,309<br>(9.3%) | 21,648<br>(9.3%) | 18,981<br>(9.3%) | 1,736<br>(8.2%) | 2,176<br>(8.6%) | 2,591<br>(9.0%) | 3,661<br>(9.1%) | 2,667<br>(9.8%) |
| **Acute upper respiratory infection** | 15,235<br>(5.6%) | 13,063<br>(5.6%) | 12,004<br>(5.9%) | 1,429<br>(6.8%) | 1,646<br>(6.5%) | 1,797<br>(6.2%) | 2,172<br>(5.4%) | 1,059<br>(3.9%) |
| **Open wound of finger(s)** | 14,781<br>(5.4%) | 14,035<br>(6.1%) | 13,181<br>(6.5%) | 272<br>(1.3%) | 345<br>(1.4%) | 439<br>(1.5%) | 746<br>(1.9%) | 854<br>(3.1%) |

**Figure 1.**
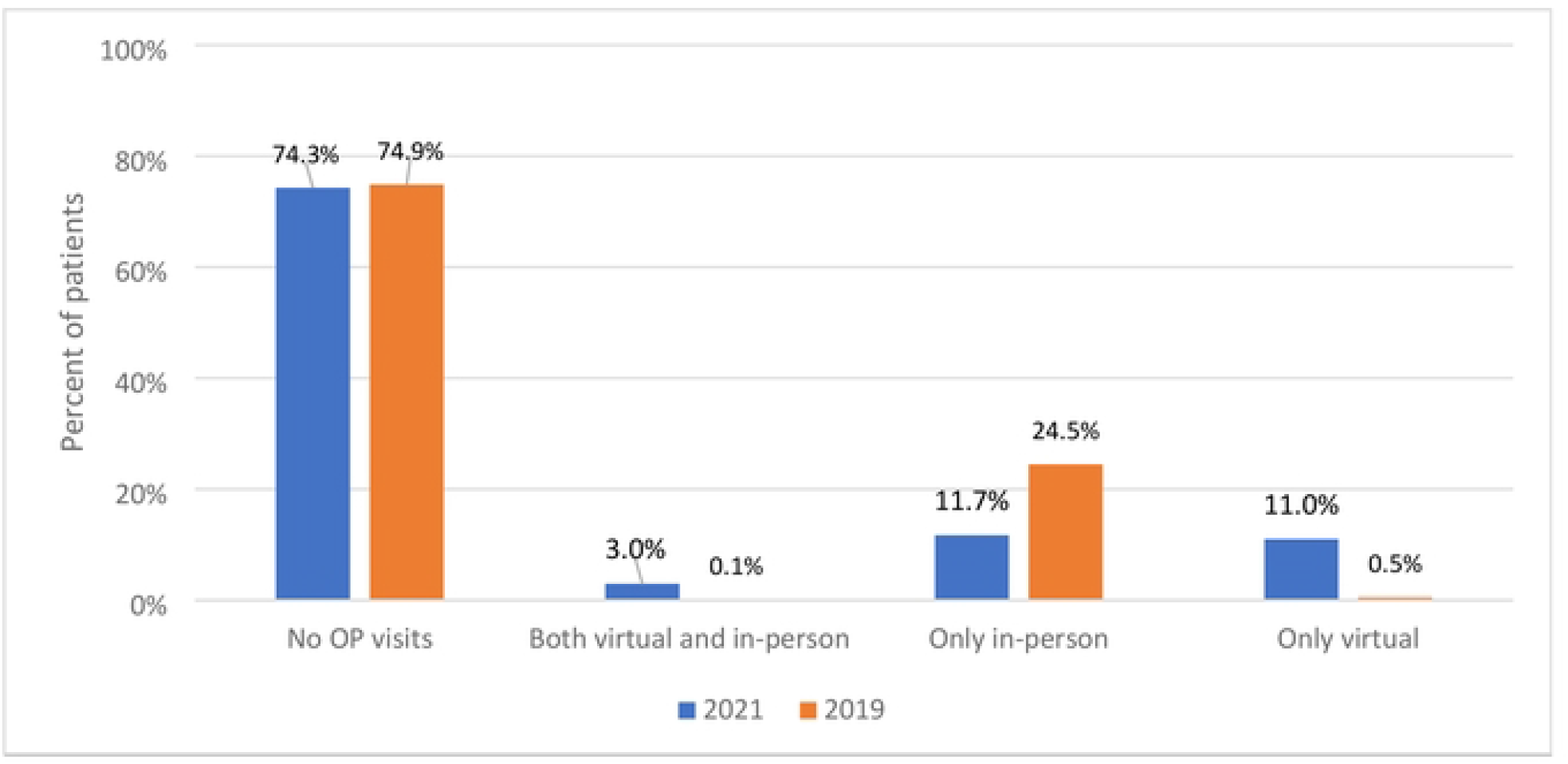
Mode of Outpatient (OP) Visits in the 7 days prior to ED admission between July 1 and Sep 30 in 2019 (n=1,113,230) and 2021 (n=1,080,334)

### Sociodemographic characteristics

Table 1 lists sociodemographic characteristics of all patients with an ED admission in 2021 (Table 1.1) and in 2019 (Table 1.2) across groups of patients with varying use of outpatient care prior to admission. When comparing the periods in 2021 vs. 2019, there were few differences in age, sex, neighborhood income, and region and rurality of residence between patients with no prior visits and patients with only in-person visits prior to their ED admission (<1% changes across). There were shifts in the characteristics of patients who used telemedicine within the 7 days prior to ED admission, with more people under 18 (12% in 2021 vs. 8% in 2019) and over 65 (30% in 2021 vs 18% in 2019) using telemedicine, more women than men used telemedicine in 2021 (59% of users were women in 2021 vs 52% in 2019). Patients living in the lowest neighborhood income regions decreased their use of telemedicine services prior to ED admission in 2021 (21% of users were in the lowest quintile vs. 29% in 2019), while those living in the highest income regions increased their use of telemedicine services (18% in 2021 vs. 14% in 2019).

Finally, patients living in urban regions, and especially in Central Ontario, increased their use of telemedicine services prior to ED admission (38% of telemedicine users were in Central Ontario in 2021 vs. 18% in 2019), while rural and North and West regions decreased their use in 2021 relative to 2019, despite absolute numbers increasing (rural: 5% in 2021 vs 13% in 2019; West 24% in 2021 vs. 32% in 2019 and North 17% in 2021 vs. 2% in 2019).

### Prior healthcare utilization

The average number of outpatient visits in the year prior to their ED admission across all patients admitted to ED in 2021 and in 2019 was similar (approximately 12 visits, Table 1). Patients with virtual visits prior to ED admission also had similar prior outpatient use in 2021 and 2019, but had more total outpatient visits relative to the entire ED user population (19 visits in 2021 vs. 21 visits in 2019). Across all patients with ED admissions, the average number of ED admissions and hospitalizations in the year prior to their current ED admission were the same in 2021 and 2019 (Mean=2.5, SD= 3.6 for ED admissions, Mean=1.5, SD= 1.2 for hospitalizations in 2021, Mean=2.5, SD= 3.6 for ED admissions, Mean=1.5, SD= 1.1 for hospitalizations in 2019, Table 1). Patients who used telemedicine prior to ED admission also had similar ED admissions and hospitalizations in 2021 and 2019 (Mean=2.7, SD=3.8 ED admissions and Mean=1.6, SD=1.2 for hospitalizations in the year prior to admission in 2021 and Mean=2.7, SD=3.8 ED admissions and Mean=1.6, SD=1.3 for hospitalizations in the year prior to admission 2019).

When examining all patients with ED admissions, the average number of outpatient visits per patient in the 7 days prior to admission did not change (Mean=1.3, SD=0.7 in both 2021 and 2019). Patients with prior telemedicine use were more likely to be hospitalized after their ED admission (13%) than patients with no prior outpatient care (7.7%), but patients with in-person visits prior to ED admissions were even more likely to be hospitalized (14.5%) (Figure 2).

**Figure 2.**
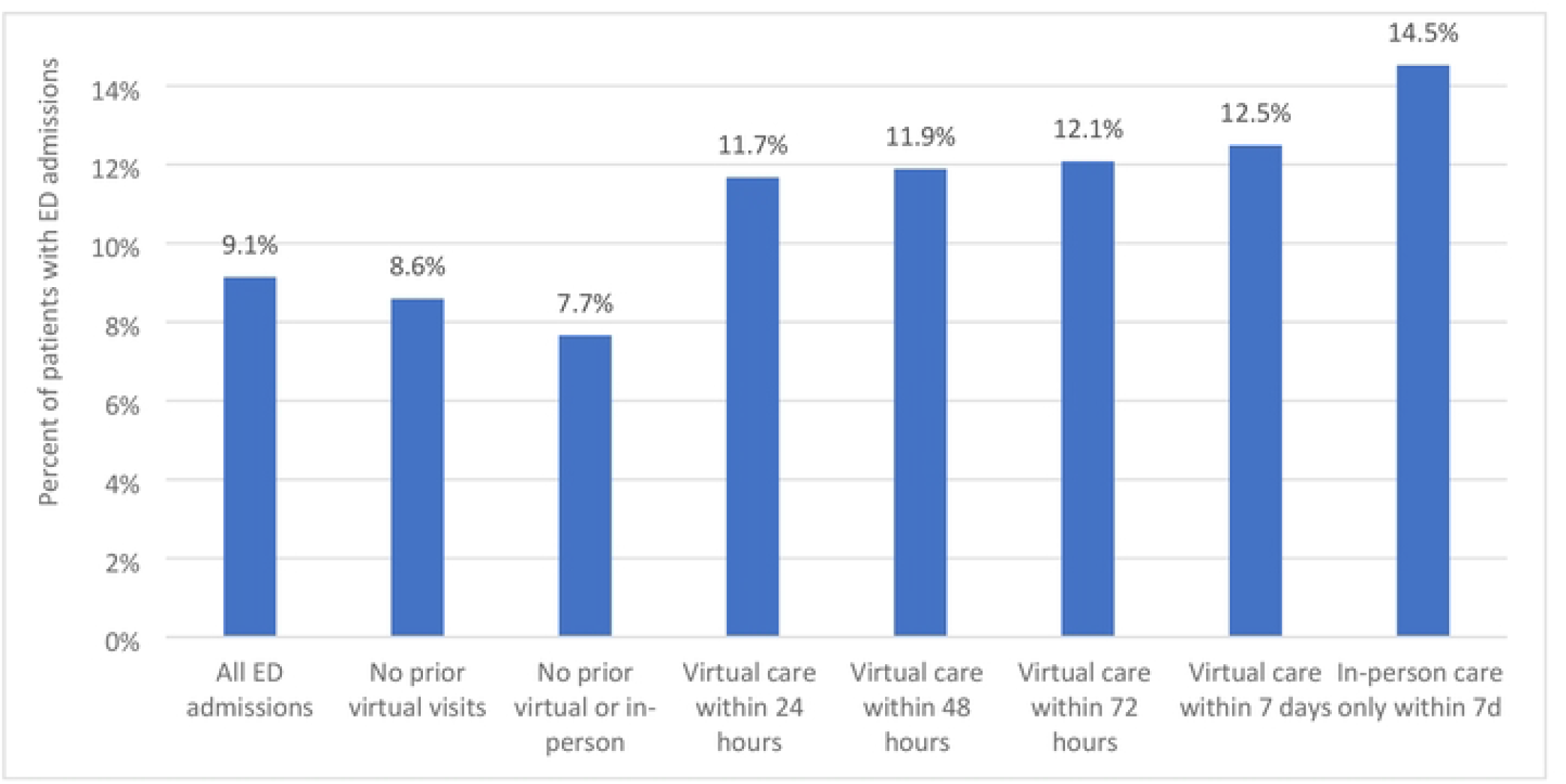
Percentage of patients with ED admissions that resulted in hospitalization across subgroups of patients with varying use of virtual care prior to admission.

### Reason for ED admission

The 5 most common reasons for ED admission in July 1, 2021-September 30, 2021 were chest pain (14%), abdominal pain (11%), urinary tract infection (9%), acute upper respiratory infection (6%) and open wound of finger(s) (5%) across all ED admission. These reasons were consistent across patient groups with little variation on volumes of these admissions (Table 2). The most common reasons for the last virtual visit prior to ED admission were “other ill-defined conditions” (12%), gastrointestinal issues (12%), anxiety (11%), chest pain (8%) and leg cramps (7%). The same reasons were seen in the in-person visits only group with the exception that gastrointestinal issues were the top reason for an in-person visits prior to ED admission (13%) (Table 3).

**Table 3:** Top 5 reasons for the last virtual visit prior to ED admission, N (%), July 1, 2021-September 30, 2021

|  | All patients | Virtual Visits within 24 hours | Virtual Visits within 48 hours | Virtual Visits within 72 hours | Virtual Visits within 7 days | In-person Visits only within 7 days |
| --- | --- | --- | --- | --- | --- | --- |
|  | N=1,080,334 | N=76,027 | N=92,749 | N=106,801 | N=151,155 | N=126,746 |
| **Other ill-defined conditions** | 8,926 (11.6%) | 4,961 (12.2%) | 5,935 (12.1%) | 6,635 (11.9%) | 8,926 (11.6%) | 4,601 (9.43%) |
| **Gastrointestinal issues** | 8,880 (11.6%) | 5,634 (13.9%) | 6,444 (13.1%) | 7,060 (12.6%) | 8,880 (11.6%) | 6,522 (13.36%) |
| **Anxiety** | 8,263 (10.8%) | 2,921 (7.2%) | 3,968 (8.1%) | 4,941 (8.8%) | 8,263 (10.8%) | 6,285 (12.88%) |
| **Chest pain** | 5,787 (7.5%) | 4,086 (10.1%) | 4,506 (9.2%) | 4,832 (8.7%) | 5,787 (7.5%) | 4,008 (8.21%) |
| **Leg cramps** | 5,178 (6.8%) | 2,755 (6.8%) | 3,313 (6.8%) | 3,774 (6.8%) | 5,178 (6.8%) | 3,723 (7.63%) |

### Physician Characteristics

There were no differences in physician characteristics (age, sex, years in practice, patient volume, and region of practice) between those who provided a virtual visit right before admission versus those who provided either a virtual or in-person visit in July 1, 2021-September 30, 2021. (Table 4)

**Table 4:** Comparison of physicians who provided the last outpatient visit before ED admissions across care modalities (virtual vs. virtual or in-person) in July 1, 2021-September 30, 2021.

|  |  | All physicians | Primary Care physicians only | Psychiatry | Pediatrics | Internal Medicine | Obstetrics & Gynaecology |
| --- | --- | --- | --- | --- | --- | --- | --- |
|  |  | N=19,704 | N=11,189 | N=1,405 | N=781 | N=763 | N=567 |
| Age Mean (SD) |  |  |  |  |  |  |  |
| Virtual Visit | Mean (SD) | 49.39 ± 12.51 | 49.09 ± 12.91 | 54.05 ± 13.40 | 50.53 ± 12.67 | 49.84 ± 13.81 | 49.31 ± 11.29 |
| Virtual or In-person visit | Mean (SD) | 49.33 ± 12.67 | 48.95 ± 13.00 | 53.56 ± 13.48 | 49.87 ± 12.48 | 48.64 ± 14.02 | 49.89 ± 11.61 |
| Sex, N (%) |  |  |  |  |  |  |  |
| Virtual Visit | Female | 8,920 (45.3%) | 5,611 (50.2%) | 615 (43.8%) | 465 (59.5%) | 226 (29.6%) | 375 (66.1%) |
|  | Male | 10,784 (54.7%) | 5,578 (49.9%) | 790 (56.2%) | 316 (40.5%) | 537 (70.4%) | 192 (33.9%) |
| Virtual or In-person visit | Female | 11,096 (42.9%) | 6,335 (48.3%) | 748 (44.3%) | 680 (59.8%) | 364 (32.2%) | 550 (64.6%) |
|  | Male | 14,791 (57.1%) | 6,777 (51.7%) | 942 (55.7%) | 458 (40.3%) | 768 (67.8%) | 302 (35.5%) |
| Region of practice, N (%) |  |  |  |  |  |  |  |
| Virtual Visit | Missing | 122 (0.6%) | 80 (0.7%) | 11 (0.8%) | 7(0.9%) | *1 – 5 | *1 – 5 |
|  | Central | 5,952 (30.2%) | 3,574 (31.9%) | 283 (20.1%) | 272 (34.8%) | 220 (28.8%) | 159 (28.0%) |
|  | East | 4,667 (23.7%) | 2,705 (24.2%) | 278 (19.8%) | 176 (22.5%) | 166 (21.8%) | 138 (24.3%) |
|  | North | 951 (4.8%) | 605 (5.4%) | 47 (3.4%) | 11 (1.4%) | *36 – 40 | *23 – 27 |
|  | Toronto | 3,049 (15.5%) | 1,446 (12.9%) | 471 (33.5%) | 141 (18.1%) | 86 (11.3%) | 103 (18.2%) |
|  | West | 4,963 (25.2%) | 2,779 (24.8%) | 315 (22.4%) | 174 (22.3%) | 250 (32.8%) | 139 (24.5%) |
| Virtual or In-person visit | Missing | 181 (0.7%) | 106 (0.8%) | 16 (1.0%) | 10 (0.9%) | 8 (0.7%) | 3 (0.4%) |
|  | Central | 7,454 (28.8%) | 3,986 (30.4%) | 335 (19.8%) | 381 (33.5%) | 327 (28.9%) | 225 (26.4%) |
|  | East | 6,268 (24.2%) | 3,220 (24.6%) | 349 (20.7%) | 253 (22.2%) | 244 (21.6%) | 224 (26.3%) |
|  | North | 1,406 (5.4%) | 846 (6.5%) | 68 (4.0%) | 32 (2.8%) | 51 (4.5%) | 38 (4.5%) |
|  | Toronto | 3,917 (15.1%) | 1,632 (12.5%) | 547 (32.4%) | 180 (15.8%) | 140 (12.4%) | 146 (17.1%) |
|  | West | 6,661 (25.7%) | 3,322 (25.3%) | 375 (22.2%) | 282 (24.8%) | 362 (32.0%) | 216 (25.4%) |
| Years in practice, mean (SD) |  |  |  |  |  |  |  |
| Virtual Visit | Mean (SD) | 22.54 ± 13.71 | 21.96 ± 14.08 | 27.20 ± 14.99 | 24.28 ± 14.06 | 22.57 ± 15.72 | 22.68 ± 12.35 |
| Virtual or In-person visit | Mean (SD) | 22.41 ± 13.78 | 21.75 ± 14.14 | 26.60 ± 15.09 | 23.45 ± 13.88 | 21.32 ± 15.92 | 23.29 ± 12.92 |
| Patient volume per day during observation window, mean (SD) |  |  |  |  |  |  |  |
| Virtual Visit | Mean (SD) | 16.15 ± 11.38 | 18.63 ± 12.43 | 6.86 ± 4.79 | 13.69 ± 10.01 | 12.16 ± 8.73 | 18.65 ± 9.27 |
| Virtual or In-person visit | Mean (SD) | 14.88 ± 11.56 | 17.36 ± 12.64 | 6.52 ± 4.79 | 11.61 ± 9.39 | 10.51 ± 8.43 | 18.06 ± 8.83 |

### Interpretation

We found no rise in ED admission volumes and pre-admission outpatient care in the summer of 2021 relative to 2019. For patients with outpatient care in the 7 days prior to admission (25% of all patients), the modality of outpatient care shifted from patients having almost exclusively in-person visits in 2019 to patients having either telemedicine only or in-person care only in the week prior to their ED admission. Very few patients (3%) had a mix of both virtual and in-person care in the 7 days before their admission. Patients who used telemedicine prior to ED admission had more outpatient visits, but similar ED and hospitalization rates in the year prior to admission. They were also more likely to be hospitalized after their ED admission than patients with no prior visits. The most common reasons for ED admission were similar across groups of patients with varying levels of telemedicine use prior to ED admission.

Early in the pandemic, there were numerous reports of a decline in ED admissions across the globe (12– 14). However, stable pandemic periods, such as the one we report on in this study, have shown a return to regular ED use (14–16). Consistent with these findings, ED admission volumes in Ontario remained nearly identical to those during a matched pre-pandemic period and the use of outpatient services prior to ED admissions did not increase. Therefore, the rise in use of virtual outpatient care in Ontario (about 50% of all outpatient care (17)) did not appear to lead to increased use of the ED. Furthermore, patients were admitted to the ED for similar reasons in both 2021 and 2019 with no evident shift in the top causes of ED admissions. To our knowledge, this is the first study to look at the relationship between telemedicine use and ED use during the pandemic.

Patients who went to the ED with prior virtual visits were more likely to be hospitalized than those without any prior outpatient care (despite similar ED and hospital utilization in the year prior). This suggests that physicians were likely able to successfully divert milder patients away from the ED through virtual visits and in most of these cases an in-person visit was not needed to make that decision (only 3% of patients had a mix of virtual and in-person visits in the week prior to ED admission). Our finding is supported by the success of numerous emergency department clinics that introduced telemedicine during the pandemic(18–20).

Telemedicine use was more prevalent in urban and higher income regions. Patients who went into the ED with a prior virtual visit in the week leading to their admission generally seemed to have better access to outpatient care, as evidenced by a higher number of outpatient visits in the year before their ED admission. While this may suggest greater severity of disease, there were no differences in the number of hospitalizations or ED admissions in the year prior to admission suggesting that the difference may lie in better access to outpatient care. This supports a growing concern that the shift towards telemedicine use may be further limiting marginalized patient populations’ access to care (21). This reinforces the notion that health equity considerations should always be at the forefront of the implementation of telemedicine programs(22,23).

As telemedicine continues to be an integral part of medicine, it is reassuring to see that despite its high use during the COVID-19 pandemic, increases in ED use were not observed. For this study we examined patients who were visiting the ED, but what remains to be examined is how telemedicine affects downstream use of not only the ED, but also other types of outpatient care and diagnostics. The balance of virtual and in-person care may shift over time, so it is important to examine the relationship between virtual outpatient use and ED use and explore how these relationships change as practices build telemedicine into their workflows and better understand its value. Telemedicine may have differential impact on various ED admission diagnoses, which can be the subject of future work. It is noteworthy that despite the ease of telemedicine visits, similarly large proportions of patients go straight to the ED without seeking outpatient care before and during COVID, likely due to poor telemedicine access. This supports the idea that we need to make the type of care available in ED or holistic care more accessible overall so that more low acuity care shifts outside the emergency department. (24).

### Limitations

Limitations of this study include the use of administrative databases which lack the clinical granularity to assess details such as the appropriateness of the visit(s). Second, the recent temporary COVID-19 virtual billing codes do not distinguish between telephone and video, and therefore we were unable to make comparisons of the various modalities of telemedicine. Lastly, the findings in this report are descriptive only and span a short period of three months, and therefore results are only preliminary and may not be generalizable in different contexts.

## Conclusions

Despite concerns that access to telemedicine may lead to a rise in ED admissions or a greater use of outpatient services prior to ED admissions (in the form of patients having both virtual and in-person visits), the net amount of ED admissions and outpatient care prior to admission remained the same over a period of the COVID-19 pandemic when cases were relatively stable. Telemedicine seems to be able to appropriately triage patients to the ED and may even prove beneficial for diverting patients away from the ED when an ED admission is not appropriate.

## Data Availability

The data is not publicly available and access is limited to the Institute for Clinical Evaluative Sciences (ICES) (https://www.ices.on.ca/), which is a prescribed entity under section 45 of Ontario’s Personal Health Information Protection Act (PHIPA). Researchers, students, policy makers or knowledge users who are affiliated with a publicly funded, not-for-profit organization and who want to obtain and analyze ICES data to answer a research question may submit a request to ICES DAS (https://www.ices.on.ca/DAS/Public-Sector). DAS staff will contact the requestor to discuss the project’s feasibility, timeline and cost. Projects requesting access to data require the approval of a research ethics board. We are happy to share our data creation plan specifying our analyses if you contact the corresponding author and in turn you can use the information in the data creation plan to run the same or follow-up analyses. A list of all datasets used is available in the paper.

## Notes

### Competing Interest Statement

The authors have declared no competing interest.

### Funding Statement

The study was funded by the Ministry of Health of Ontario, Canada.

### Author Declarations

Use of these databases for the purposes of this study was authorized under §45 of Ontario’s Personal Health Information Protection Act, which does not require review by a research ethics board (REB). All data was de-identified at ICES and individual patient consent was waived.

